# Levels of the TNF related cytokine, LIGHT increase in hospitalized COVID-19 patients with Cytokine Release Syndrome and ARDS

**DOI:** 10.1101/2020.07.27.20152892

**Authors:** David S. Perlin, Inbal Zafir-Lavie, Lori Roadcap, Shane Raines, Carl F. Ware, Garry A. Neil

## Abstract

Many COVID-19 patients demonstrate lethal respiratory complications caused by cytokine release syndrome (CRS). Multiple cytokines have been implicated in CRS, but TNFSF14 (LIGHT) has not been previously measured in this setting. In this study, we observed significantly elevated serum LIGHT levels in hospitalized COVID-19 patients as compared to healthy age and gender matched control patients. The assay detected bioavailable LIGHT unbound to the inhibitor Decoy receptor-3 (DcR3). Bioavailable LIGHT levels were elevated in patients both on and off ventilatory support, with a trend toward higher levels in patients requiring mechanical ventilation. In hospitalized patients over the age of 60, who exhibited a mortality rate of 82%, LIGHT levels were significantly higher (p=0.0209) in those who died compared to survivors. As previously reported, interleukin 6 (IL-6) levels were also elevated in these patients with significantly (p=0.0076) higher levels observed in patients who died vs. survivors, paralleling the LIGHT levels. Although attempts to block IL-6 binding to its receptor have shown limited effect in COVID-19 CRS, neutralization of LIGHT may prove to be more effective owing to its more central role in regulating antiviral immune responses.

---

The findings presented herein demonstrate that LIGHT is a cytokine which may play an important role in COVID-19 patients presenting ARDS and CRS and suggest that LIGHT neutralization may be beneficial to COVID-19 patients.

A leading cause of death in patients infected with SARS-CoV-2 (COVID-19 patients) is an unregulated immune response to the virus and/or resulting cell damage. The excess release of inflammatory cytokines can be initiated by the death of infected cells known as pyroptosis (1). Pyroptosis is known to drive the release of pro-inflammatory cytokines, attracting inflammatory cells to the site of infection and generating a positive feedback loop of cytokine secretion. This excess secretion of cytokines results in an ineffective and exaggerated immune response coined cytokine release syndrome (CRS), which also occurred in SARS-CoV and MERS-CoV (2, 3). The clinical consequence of CRS in COVID19 is acute respiratory disease syndrome (ARDS) and associated complications, which results low blood oxygen levels therefore requiring supplemental oxygen therapy and mechanical ventilation. ARDS accounts for the cause of 70% of COVID-19 death cases (4). Despite intensive support, patients often progress to fatal respiratory failure and/or multi-organ complications including heart, liver and renal failure which account for 28% of lethal cases (5, 6).

Several specific cytokines and immunomodulators have been observed in the serum/plasma of COVID-19 patients and postulated to be the driving force for the uncontrolled immune response. Among these are IL-6, IP-10, MCP-1, IL-1β (7). Indeed, symptomatic COVID19 patients show elevation of several cytokines (TRAIL, MCSF, GRO-α, GCSF and IL6, among others) compared to asymptomatic persons (8). As a result, several potentially promising cytokine neutralizing therapies have reached clinical trials. While dexamethasone therapy appears to be effective in some patients, robust clinical responses have not been consistently observed (7). There remains an urgent need to find novel targeted therapies for CRS and ARDS in COVID-19.

The TNF-related cytokine LIGHT (*TNFSF14*) has proinflammatory activity with multifaced roles in stimulating T-cells and innate immune responses (9). LIGHT engages two cellular signaling receptors, Lymphotoxin β receptor LTβR and HVEM (TNFRSF14) and is inactivated by decoy receptor-3 (DcR3). Circulating bioavailable LIGHT unbound to DcR3, “free LIGHT”, is implicated as a pathogenic cytokine in viral airway infections. In addition, LIGHT is a key factor in rhinovirus-induced chronic lung inflammation (10) and increased in neutrophils and macrophages from patients with viral pneumonia due to adenovirus 55 infection (11). LIGHT is also known to be involved in orchestrating uncontrolled immune responses resulting in autoimmunity and tissue injury such as Inflammatory Bowel Disease (IBD), asthma and lung fibrosis (12). Additionally, LIGHT induces the release other inflammatory cytokines including IL-6 and GM-CSF (13), which are reported at elevated levels in COVID-19 patients.

To better understand the potential importance of LIGHT in the mechanism of host response to the virus, we analyzed the serum of 47 COVID-19 patients hospitalized at the Hackensack Meridian Health Hospital, by measuring serum IL-6 and free LIGHT using a validated immunoassay, performed by Myriad RBM.

The data provides compelling evidence that hospitalized patients diagnosed with COVID-19, both on and off ventilatory support, have significantly higher free LIGHT levels than healthy age- and gender-matched controls (Figure 1A). The elevated serum levels of free LIGHT are within the receptor signaling concentrations, and likely exceed the sequestering action of DcR3. In hospitalized patients over the age of 60 who exhibited a mortality rate of 82%, free LIGHT was significantly higher in those who died compared to those who survived (Figure 1B). As previously reported (14) IL-6 was also elevated in COVID-19 infected patients. We observed that the highest IL-6 levels were detected in ventilated patients (Figure 1C). Similar to LIGHT hospitalized patients over the age of 60 who died had higher IL-6 levels than patients who recovered (Figure 1D).

**Figure 1:**
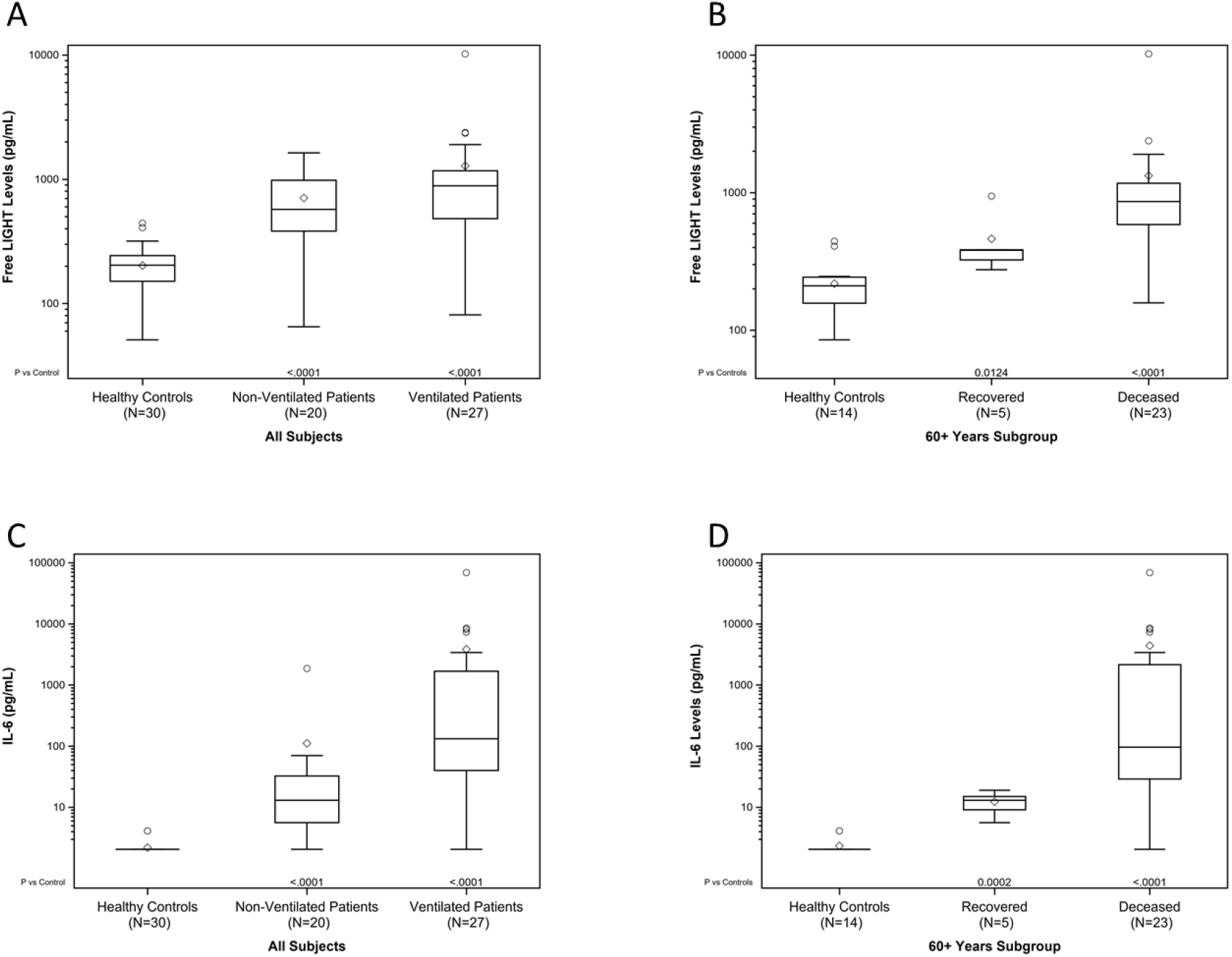
IL-6 and Free LIGHT levels in serum of COVID-19 patients. **(A)** Free LIGHT serum levels in COVID-19 ventilated (n=27) and non-ventilated (n-=20) COVID-19 patients compared to healthy controls (n=30). **(B)** Free LIGHT serum levels in COVID-19 patients at the age of 60 years old or older, grouped by clinical outcome (recovered vs. deceased). **(C)** IL-6 serum levels in COVID-19 ventilated (n=27) and non-ventilated (n-=20) COVID-19 patients compared to healthy controls (n=30). **(D)** IL-6 serum levels in COVID-19 patients at the age of 60 years old or older, grouped by clinical outcome (recovered vs. deceased). IL-6 was measured using a validated immunoassay on the Luminex platform. Free LIGHT was measured using a validated immunoassay on the Quanterix™ ultra-high sensitive SIMOA® platform. P value was calculated using the Kruskal-Wallis test.

LIGHT is an important and central regulator of the immune response in barrier tissues, including the upper respiratory tract and lung (15).

LIGHT’s actions are mediated by two receptors, HVEM and LTβR (16). The LTβR is expressed in macrophages, neutrophils, stromal cells and epithelial cells, whereas HVEM is also expressed in T and B lymphocytes. LIGHT induces the release of tissue damaging inflammatory cytokines, including IL-6, via HVEM, and promotes high endothelial cell activation allowing inflammatory cell accumulation at sites of virus infection through LTβR. LIGHT’s effects are modulated by two mechanisms: a circulating decoy receptor, DcR3 that limits the bioavailability of LIGHT to both receptors, and the inhibitory checkpoint molecule, BTLA that controls HVEM activation (16). We hypothesize that high levels of LIGHT induced in some patients with COVID-19 pneumonia, may overwhelm DcR3 and BTLA resulting in unregulated activation of HVEM and LTβR and CRS. The observation that free LIGHT levels are, indeed, elevated in these patients and correlate to some extent with clinical outcomes supports this hypothesis.

Along with vaccines and anti-viral therapies, there remains an urgent need for therapies for CRS and ARDS in COVID-19 infected patients. LIGHT’s role as an immune modulator and a driver of inflammatory response and its presence in the serum of COVID-19 patients makes it a plausible target for intervention. We have therefore initiated a clinical trial of CERC-002, a novel neutralizing human anti-LIGHT monoclonal antibody in the COVID-19 patients with early ARDS in the US to test this hypothesis (NCT04412057).

## Methods

### Measurement of Free LIGHT

Free LIGHT assays were performed by Myriad RBM in their central laboratory in Austin, TX, using Quanterix’s fully automated HD-1 Analyzer and single molecule array (Simoa) technology as described elsewhere(17, 18). All incubations were performed at room temperature inside the Simoa HD-1 analyzer. Capture antibody conjugated paramagnetic beads were incubated with standards, samples or controls and biotinylated detection antibodies. The beads were then washed and incubated with streptavidin-ß-galactosidase (SßG). After the final wash, the beads were loaded into the Simoa Disc with enzyme substrate, resorufin ß-galactopyranoside (RGP). The fluorescence signals are compared to the standard curve and the quantity of free LIGHT was determined for each sample. The lower and upper limits of detection for free LIGHT were determined to be 0.8 – 4,000 pg/ml.

### Measurement of IL-6

IL-6 levels were determined as described elsewhere(18, 19) using MyriadRBM at their central laboratory in Austin, TX using their proprietary InflammationMAP™ Panel, a set of multiplexed assays which is microsphere-based and consists of using antigen-specific antibodies optimized in a capture-sandwich format. All incubations were performed at room temperature. In brief, 5 μL of a diluted mixture of capture-antibody microspheres was mixed with 5 μL blocker and 10 μL standard, pre-diluted sample, or control in a hard-bottom microtiter plate. Serum samples were diluted to the appropriate dilution. The plates were incubated for 1 hour. 10 μL biotinylated detection antibody was added to each well, thoroughly-mixed, and incubated for 1 hour. 10 μL diluted Streptavidin-phycoerythrin was then added to each well, thoroughly mixed, and incubated for 60 minutes. A filter-membrane microtiter plate was pre-wetted by adding 100 μL wash buffer followed by aspiration via a vacuum manifold device. The reaction contents of the hard-bottom plate were then transferred to the respective wells of the filter plate. All wells were vacuum aspirated and the contents were washed twice with 100 μL wash buffer. After the last wash, 100 μL wash buffer was added to each well, and the washed microspheres are resuspended with thorough mixing. The plate was then analyzed on the Luminex platform. The lower limits of detection for IL-6 was determined to be 1.4 pg/mL.

## Data Availability

N/A data was generated in this study

